# 3D-Mapping of QRS Loops: Visualizing the Ventricular Electrical Activities

**DOI:** 10.1101/2025.02.06.25321576

**Authors:** Tania Ghosal, Subhankar Kumar, Imran Ahmed, Damodar Prasad Goswami, Anupam Bandyopadhyay, Arnab Sengupta

## Abstract

This paper presents innovative visualization tools for the analysis and interpretation of three-dimensional vectorcardiogram (VCG), focusing on the QRS complex of the cardiac cycle. Traditional electrocardiography (ECG) lacks the spatial detail necessary for comprehensive cardiac evaluations; however, VCG provides a three-dimensional representation of electrical activity in the heart, allowing for a nuanced understanding of cardiac dynamics. We propose five distinct methodologies for representing spatial QRS VCG loops: (1) unmodified spatial VCG loops that track the progression of ventricular depolarization, (2) fixed-scale VCG loops that facilitate direct comparisons across individuals and conditions, (3) orientation of QRS loop with respect to three orthogonal planes in the 3D space. (4) octant-specific plots that offer insights into spatial distributions, and (5) unit vector and unit sphere representations that emphasize directional movement while normalizing magnitude. Each method has unique strengths in elucidating ventricular electrodynamics in normal and in cardiac conditions such as anterior wall myocardial infarction and inferior wall myocardial infarction, highlighting differences in loop size, directionality of propagation, orientation, and morphology. Together, these methodologies provide a robust framework for advancing VCG research and enhancing clinical assessments of cardiac function. Preliminary findings highlight the potential of these innovative tools.

## Introduction

As a complex electro-mechanical structure, the cardiac electrical activities are occasionally evaluated by vector analysis in three-dimensional space [1, 2]. It is especially relevant in certain disease conditions like myocardial infarction [3], where there is a loss of structural and functional integrity of the heart accompanied by depolarization and conduction anomalies. [4]

Cardiac vectors are fundamentally time-varying current dipoles propagating in the heart [5]. This stereotypical pattern of cardiac vector propagation can be recorded as body surface potential by specific electrode positions, and is represented as Electrocardiogram (ECG), Vectorcardiogram (VCG) etc.

While ECG represents the scalar magnitude of the component of the main electrical vectors along a particular lead axis, VCG graphically represents the trajectory of the instantaneous vector tips originating from a zero-reference point in three-dimensional space [1, 6], and provides a spatial depiction of electrical vectors, captures both the direction and magnitude of the heart’s depolarization and repolarization processes, offering detailed insights into the spatial orientation of electrical currents within the heart. It is more informative and sensitive than ECG [2, 7]. as an evaluation tool for assessing the dynamics of cardiac physiology, because of its additional degree of freedom [5, 8].

Although the VCG was discovered long ago [9, 10], the complexity, imprecision, and tedious process of constructing a VCG loop, together with the lack of established methodologies and standardisation have thus far restricted the progress of VCG research. However, the growing realization that spatial analysis of the cardiac electrical phenomena can provide information not provided by the traditionally recorded ECG has spurred renewed interest in vectorcardiogram analysis in recent years [11, 12]. Moreover, the development and widespread availability of computer-based technology, signal acquisition systems, mathematical and computational tools have simplified the construction and objective implementation of VCGs.

A number of recent articles have attempted to describe the static and dynamic morphology of the QRS loop of VCG in various conditions [13-20]

However, there exists a multiplicity of approaches and presentation patterns of spatial VCG loop; including visual feature extraction, characteristics description and calculation of different objective variables in health and in a number of disease states [3, 7, 10, 12, 14, 15, 18]. Also, correlation studies, sensitivity and specificity analysis with respect to various diagnostic and prognostic criteria, evaluation of ROC curves have been attempted. But, to date there has been no conformity towards development of normative data ranges and qualitative descriptor and there is inadequate standardisation and multiple approach of reference system, axes, scaling, calibration etc [7, 12].

As VCG signals are highly informative and sensitive, they reflect complex and detailed interactions between components, leading to emergent behaviours that are sometimes hard to standardize to a consistent visual pattern or a range of normative datasets. They can be affected by subtle changes in their environment, even with a minor mathematical transformation, making it challenging to control and standardize conditions. Being non-linear system, a small change can lead to large and disproportionate effects, and dependence on specific contexts or initial conditions makes standardization more difficult. Additionally, accurate measurement, sophisticated processing (which can be sensitive to algorithmic and parameter choices), quantification of VCG signal present considerable challenges.

Our group has been working on characterization of static and dynamic spatial VCG loops [5, 6, 8, 21], and has reported some interesting findings on loop morphology and dynamics. We believe there is ample scope to explore the descriptive spatial morphology of loops in normal persons and in various diseases that can affect the cardiac vectors. Detailed visual description of the loop and recognition of existing and emerging patterns can provide valuable information regarding the changing direction and magnitude of the serial cardiac vectors and their significance in altered morphologies associated with disease. The visual comparison and correlation of loop morphology with clinical conditions may offer important and valuable insight into the pathophysiology of disease states.

In the present paper, we intend to present a battery of effective visual tools for representing spatial QRS VCG loop for descriptive as well as analytical evaluation. We will also use these tools to illustrate examples from control volunteers and cases of acute myocardial infarction (AMI) and its subgroups of anterior wall myocardial infarction (AWMI), and inferior wall myocardial infarction (IWMI).

### Brief description of the pattern variables

#### 1. Unmodified spatial VCG loop

In order to evaluate the morphological details of the spatial VCG, each loop is constructed with a minimal scale of the reference axes frame, so that the spatial features like shape, size contour remain clearly distinguishable. Each loop is color coded to visualize the temporal order of appearance of the cardiac vectors, and spatial direction of the loop.

#### 2. Spatial VCG loop with fixed scale of axes

As the VCG loop morphology is not scale invariant, to compare the shape and size of the loop structure between different individuals, or the same individual during different stages of illness, all three axes are fixed to their maximum and minimum, enabling comparison. The color coding as described above helps to visualize the temporal order and spatial direction of the loop. Additionally, the point of origin of the loop, and the maximum spatial QRS vector is obtained, to evaluate the overall movement of the loop with respect to its origin.

#### 3. Orientation of QRS loop in orthogonal planes

The orientation of QRS loop with respect to three orthogonal planes; Frontal, Sagittal and Transverse is represented in the 3D space. Each plane is differently colour coded.

#### 4. Spatial VCG loop with octane wise distribution

The distribution pattern of the loop in various cartesian octants is also studied in order to evaluate the preferential occupation of the QRS loop in a particular quadrant or a combination of more than one quadrant in controls and also in AMI and its subgroups. The portion of the loop in each octant is colour-coded depicting the pattern of distribution in different octants. This provides valuable and objective insight into the proportionate occupation of the VCG loop in the spatial segments of cartesian octants.

#### 5. VCG Unit Vector and Unit Sphere System

Vectorcardiogram is constructed as a space curve by joining the tips of the serial instantaneous vectors with respect to a zero-reference point, according to direction, magnitude and polarity of the vectors. Accordingly, the spatial orientation of the vectors depends upon both directions and magnitudes. Smaller movement of the spatial points occurs in regions where the rate of change of magnitude of the serial vectors is less; whereas larger changes in the magnitude of the serial instantaneous vectors (e.g. the VCG loop that corresponds to the R wave of ECG) would apparently magnify the deviation of the vector orientation.

To standardize VCG representation, the spatial VCG can be transformed onto a unit sphere, using unit vectors. A unit sphere is a sphere of unit radius, where the set of spatial points is normalized at unit distance from the centre point in three-dimensional space. Here, direction component is isolated from magnitude. This allows consistent comparison and analysis of different VCG loops, with respect to the direction of the vectors and orientation of the loop.

The unit sphere serves as a universal reference frame, facilitating effective comparison and analysis across various datasets. By visualizing data points on this standardized sphere, we can discern spatial patterns, correlations, and anomalies more accurately, thereby enhancing the depth and reliability of spatial analysis outcomes.

This construction facilitates us to study the effect of varying direction, keeping aside the effect of magnitude.

This spherical framework provides valuable insights into the spatial distribution and geometrical relationships of these vectors.

The unit sphere system of VCG is also represented as a unit vector system, visualized as a space curve, without explicitly displaying the sphere.

### Methodology and visualization

#### Data Collection

Vectorcardiography data were primarily sourced from electrocardiography recordings of control volunteers and cases of myocardial infarction, obtained using a digital electrocardiograph (Biopac Instruments, USA; sampled at 1000 samples/s). For construction of Spatial vectorcardiograms, we used simultaneously recorded data of QRS complex from three orthogonal leads (I, V2 and aVF), as explained in earlier studies [8, 21].

Before the study, approval was obtained from the Institutional Ethics Committee of IPGME&R, Kolkata, India (Ref. No. IPGME&R/IEC/2021/114) and each individual subject gave their informed consent. We closely adhered to the World Medical Association’s Declaration of Helsinki, which outlines the ethical guidelines for medical research involving human participants, during the entire research project.

#### Selection of axes

In contrast to the conventional right-handed Cartesian coordinate system used by computers, the human heart’s internal electrical activity adheres to a left-handed coordinate system, due to its anatomical location on the left side of the chest. To facilitate the visualization and tracking of heart vectors more intuitively, a specialized Cartesian coordinate system has been devised for analysis. Within this custom framework, lead L1 corresponds to the y-axis, lead V2 to the x-axis, and the negative of lead AVF to the z-axis.

A Python script was created to systematically analyze the 3D spatial data. The script uses Matplotlib’s 3D plotting features to visually represent the points in 3D space.

### Data normalization process for Unit Sphere and Unit Vector analysis

Each coordinate (x, y, z) is normalized to ensure the magnitude equals 1, effectively mapping it to the surface of a unit sphere centred at the origin (0, 0, 0). This normalization ensures that all data points are proportional and comparable, allowing for accurate spatial analysis. By dividing each coordinate by its Euclidean norm, all points lie precisely on the unit sphere’s surface.

### Mapping as Unit vector and on the Unit Sphere

The spatial data were normalized and then plotted as unit vectors on a unit sphere. This visualization provides a clear way to observe the spatial relationships among the data points. The plot of unit vectors allows us to compare the plot image with the unmodified and fixed scale of axes spatial VCG loop and can precisely evaluate the direction of vector propagation and orientation of the loop. By mapping everything to a unit sphere, we can easily compare datasets and identify patterns, clusters, or anomalies.

### Visualization Process

#### 1. Unmodified form

The color-coded 3D scatter plots were generated to visualize the data points over time, and the spatial features like shape and contour. The dataset was divided into three phases: the early phase was coloured green, the mid-phase was orange, and the last phase was red. This enabled us to track the progression of ventricular depolarization and observe where the early, major, and late ventricular vectors are located.

#### 2. Modified loop with fixed scale of axes

The same space curves as above were generated, but with fixed maximum and minimum scale of axes, to enable comparison of the size of the QRS loop, between the groups and subgroups.

#### 3. Loop pattern with respect to orthogonal planes

The same space curve representing the planar orientation of the QRS loops along with depiction of the major ventricular vector were generated. Colour coding was utilized to illustrate different planes separately.

#### 4. Octant wise distribution plots

Distribution pattern of the VCG loop in various octants was represented as a 3D plot. These plots highlight the points in each octant and show the path of movement using distinct colours, along with coloured lines representing the major planes (xy, yz, and zx). The origin and the farthest point are emphasized for reference. This plot offers a comprehensive view of the spatial distribution.

#### 5. Unit sphere visualization and insights

The unit sphere visualization helps researchers observe how the normalized data points are distributed, making it easier to spot patterns, clusters, or outliers. By normalizing the data to the unit sphere, we ensure that comparisons across different datasets are fair and consistent. The 3D visualization allows for intuitive exploration, encouraging deeper insights and the development of new hypotheses.

To compare results, QRS loops from several myocardial infarction cases were plotted alongside those from the control group, using the same parameters described above.

Here we illustrate the various forms of representation of VCG loops of control volunteers, and patients with AWMI and IWMI of two illustrative subjects from each subgroup.

All the spatial points are color coded with initial, mid and terminal one third of the data points in green, orange and red respectively (Figs.1–5). These data points are expected to contain the initial septal, middle major and later terminal ventricular vectors.

Fig. 1 shows unmodified spatial QRS loops, with a minimal scale of the reference axes frame in each individual, exhibiting distinct spatial features. The illustrated images evidently distinguish between spatial features of VCG loop between control volunteers and AMI, and also within subgroups of AMI. Fig. 2 represents the spatial QRS loop constructed with fixed scale of axes and demonstrates the reduced size of the loop in cases of AMI, as compared to controls. Fig. 3 shows the spatial QRS Loop of fixed frame of reference separated by Frontal (green), Sagittal (pink) & Transverse (white) planes. There are differences in the loop orientation with reference to the said planes between the control and cases and also within the case subgroups. Also, the major ventricular vector is represented with respect to orthogonal cartesian octants by the broken pink line in each QRS loop. There is apparent difference in the octant occupation pattern along with alteration in the direction and magnitude of the major ventricular vector between groups and subgroups. The octant occupation pattern was further assessed in the Fig. 4, which exhibits the spatial QRS Loop in various cartesian octants of fixed frame of reference. The loops are colour coded with eight different colours with reference to their occupation in various octants. The difference in octant occupation pattern is clearly noticeable between the control and cases and also within the case subgroups. Fig. 5 shows the VCG Unit Sphere of a single QRS loop while Fig. 6 shows the VCG Unit Vectors of a single QRS loop for the same subjects. The unit sphere and unit vector representations elaborately depict the directional orientation of the each instantaneous QRS vector. A careful visual comparison between control and AMI cases and also between case subgroups, exhibits the pattern of alteration in accordance with the type of infarction. While for control group, the loop structure is preserved with magnitude normalization (Fig. 5, 6), in various form of AMI, loop structure discernibly altered and fragmented after magnitude correction.

**Fig. 1:**
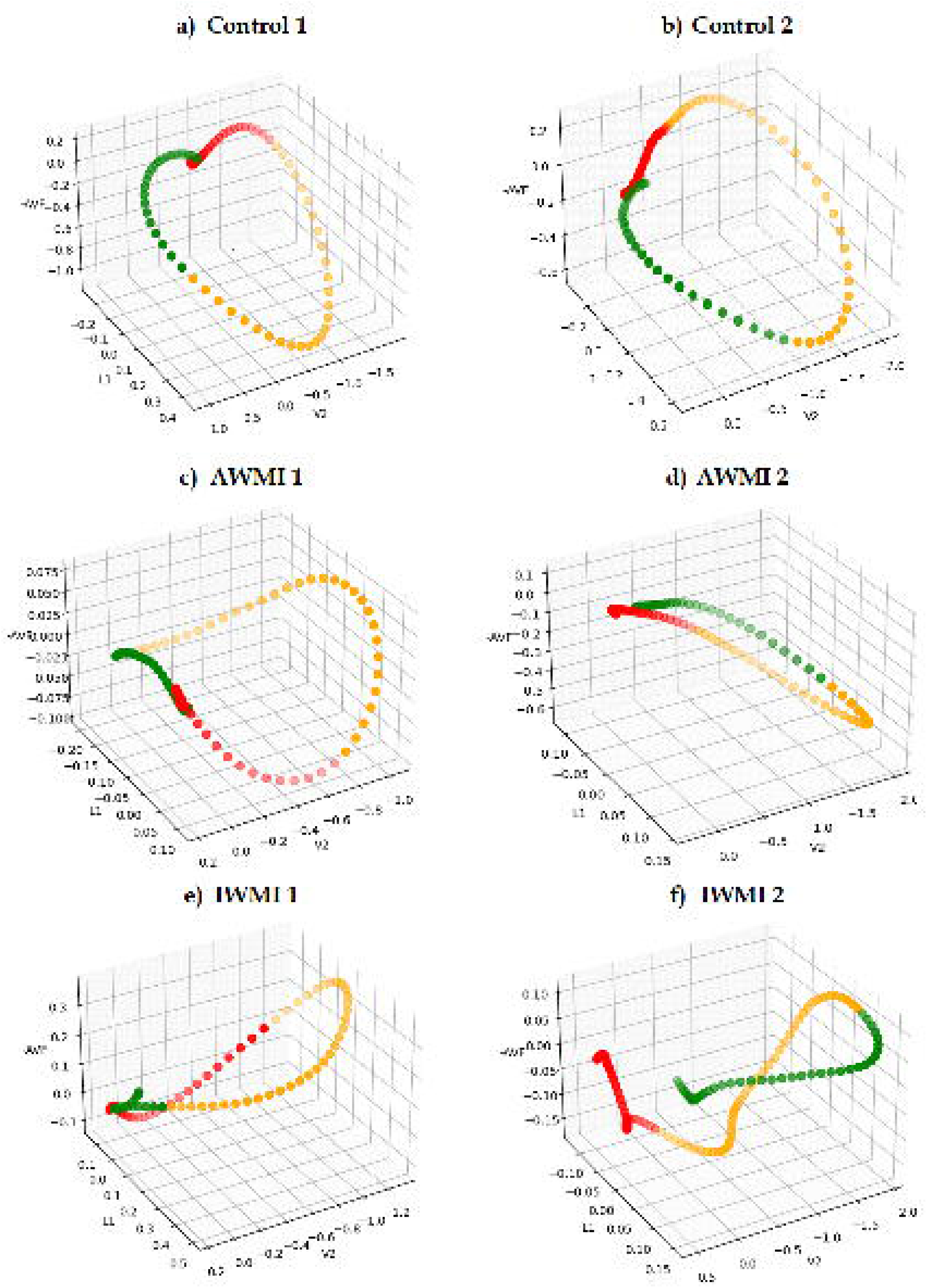
Illustrative depiction of unmodified Spatial QRS Loops in Control volunteetrs, AWMI and IWMI Patients. The loops are divided into three phases: the early phase is marked in green, the mid-phase in orange, and the final phase in red.

**Fig. 2:**
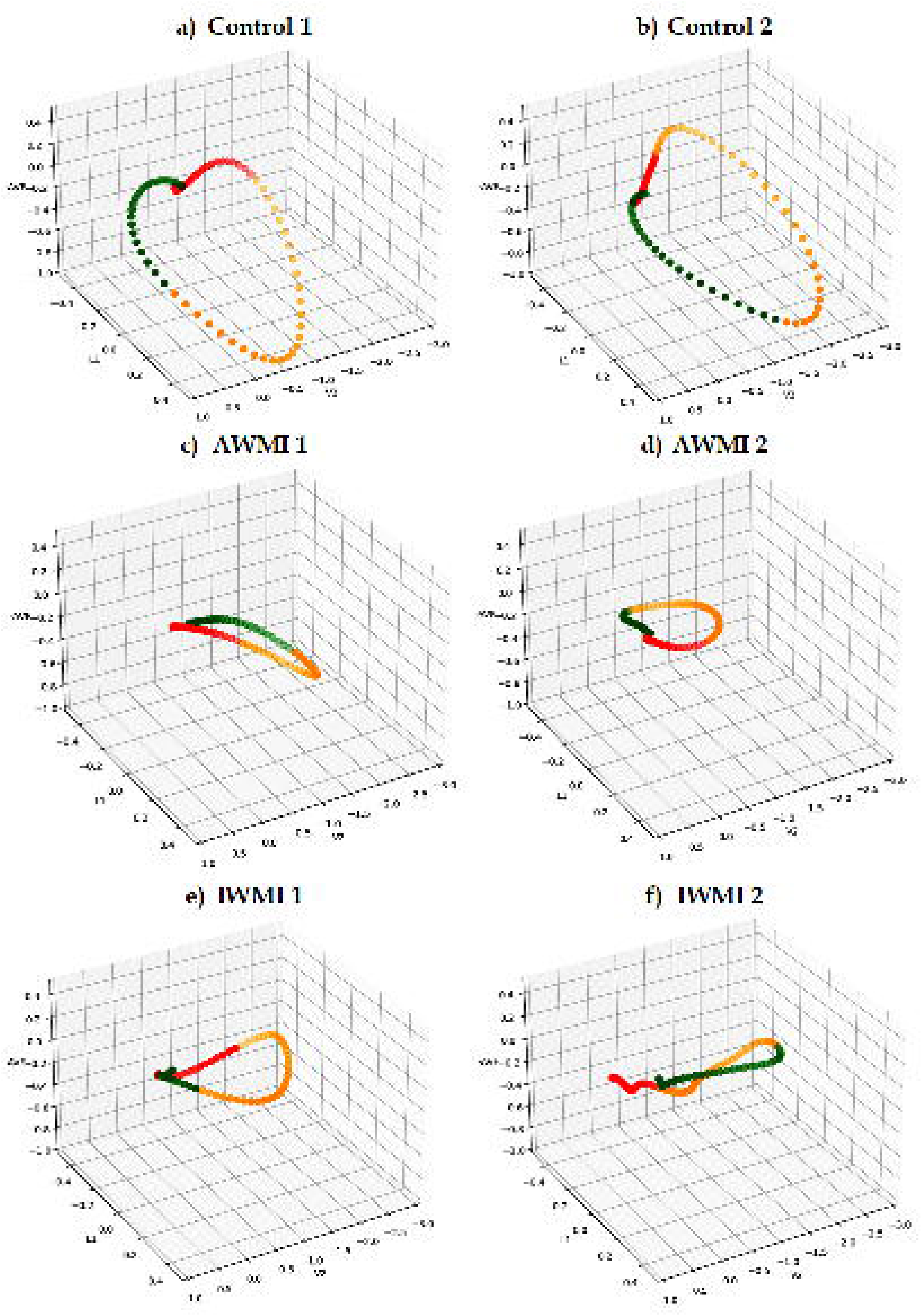
Comparative illustration of Spatial QRS Loop with fixed scale of axes in Control volunteetrs, AWMI and IWMI Patients. The loops are divided into three phases: the early phase is marked in green, the mid-phase in orange, and the final phase in red.

**Fig. 3:**
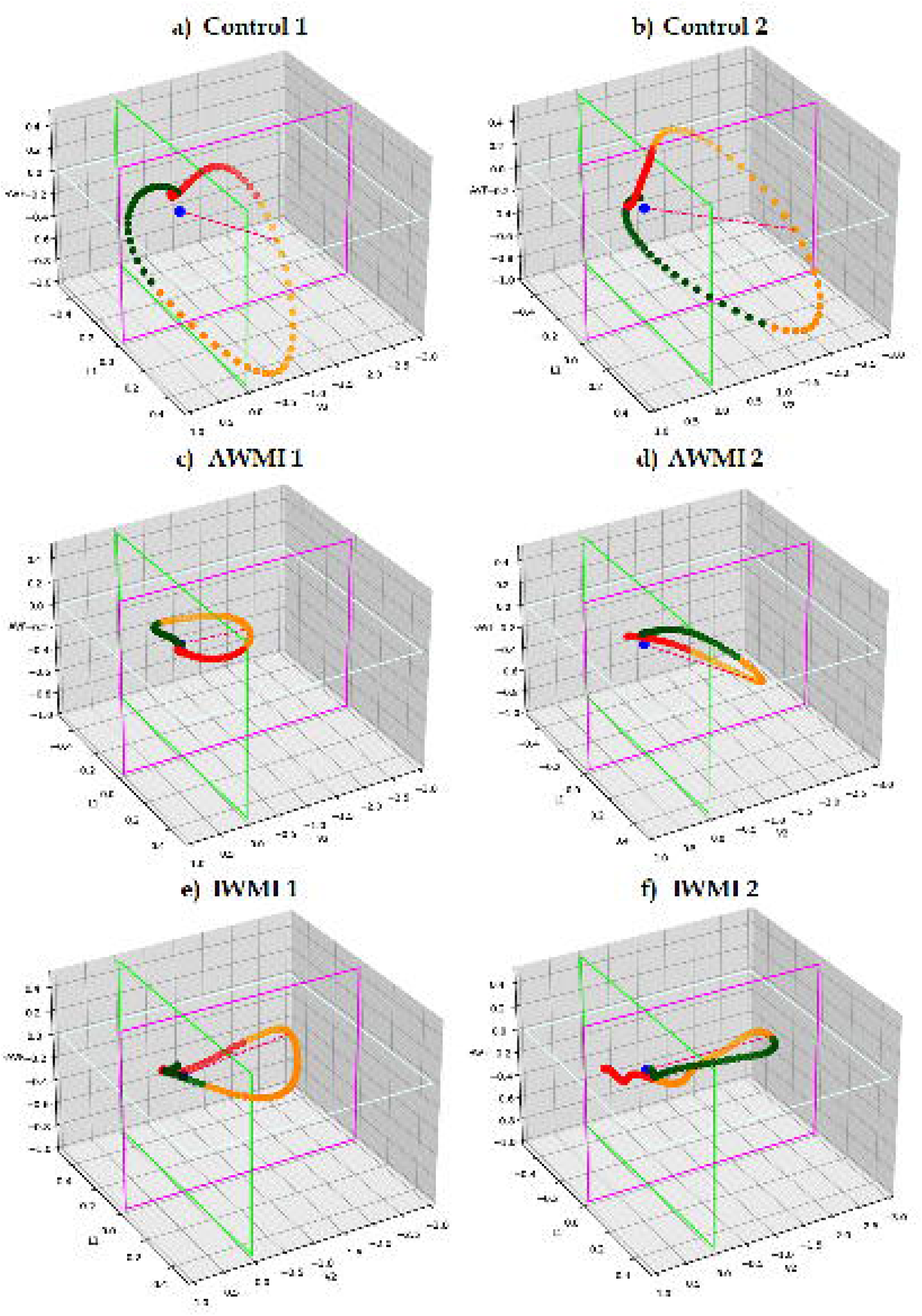
Comparative illustration of orientation of Spatial QRS Loop with respect to three orthogonal planes of fixed frame of reference separated by Frontal (green), Sagital (pink) & Transverse (white) planes in Control volunteers, Patients of AWMI and IWMI with two subjects in each group. The major ventricular vector is represented as a broken pink line. The blue dot indicates the point of origin. The loops are divided into three phases: the early phase is marked in green, the mid-phase in orange, and the final phase in red.

**Fig. 4:**
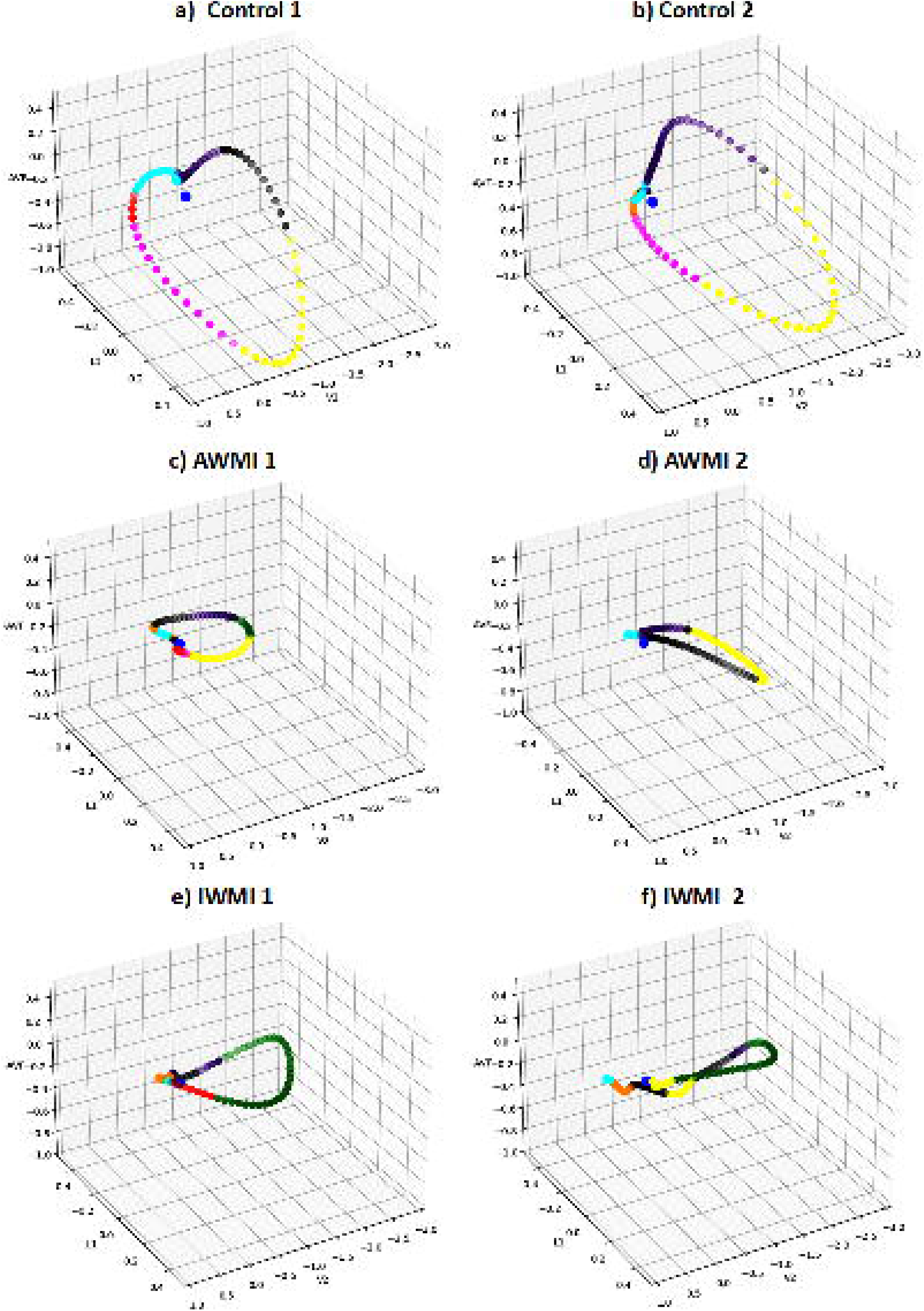
Comparative illustration of Spatial VCG Loop in cartesian octants of fixed frame of reference, depicted by various octant specific colour code of the portion of the loop in Control volunteetrs, Patients of AWMI and IWMI with two subjects in each group. The blue dot indicates the point of origin. Octant No. and Colour code 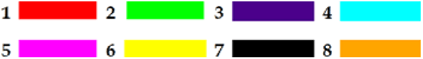

**Fig. 5:**
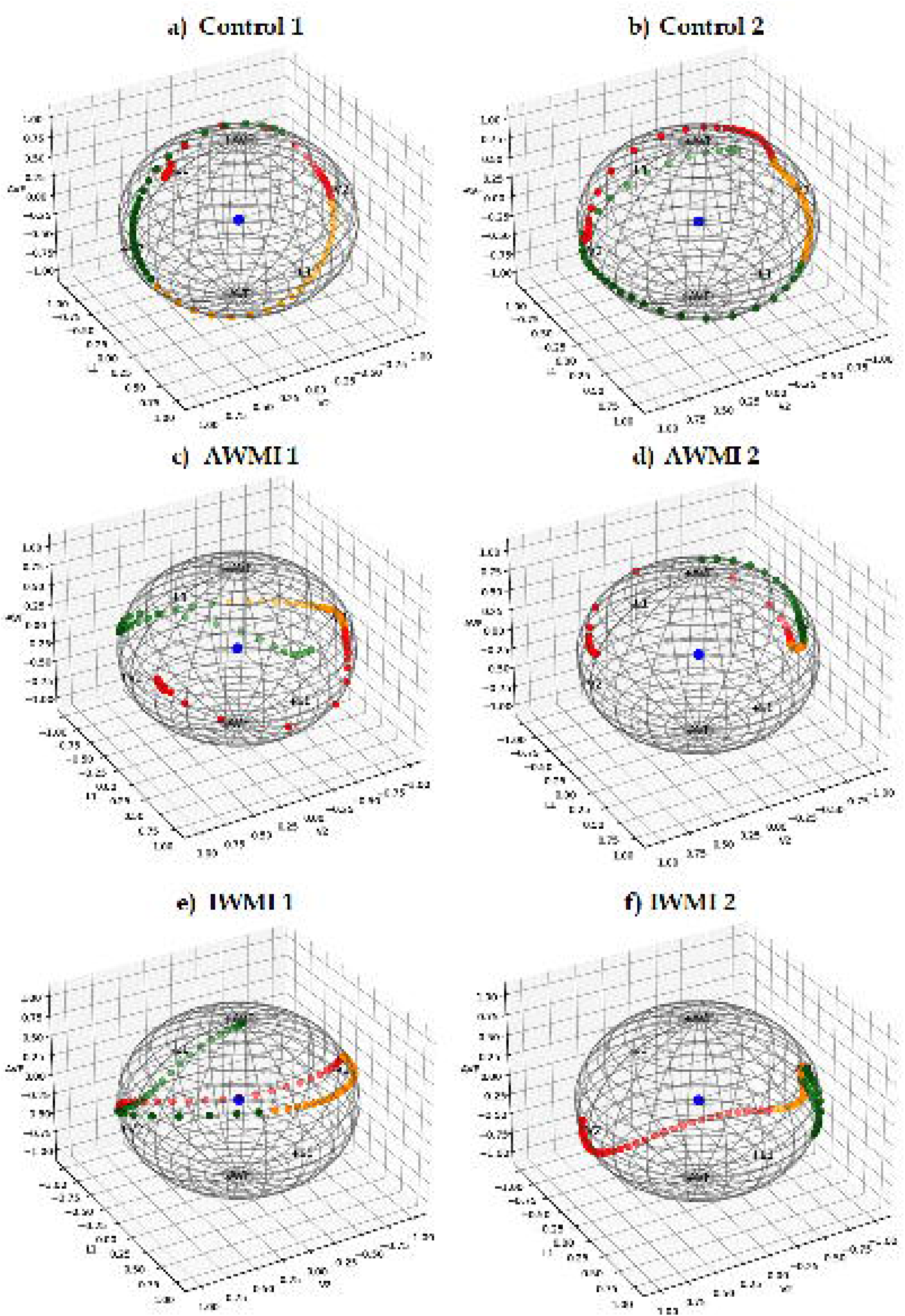
Illustrative representation of Spatial QRS Loop depicted in unit sphere system in Control volunteetrs, Patients of AWMI and IWMI with two subjects in each group. The loops are divided into three phases: the early phase is marked in green, the mid-phase in orange, and the final phase in red. The blue dot indicates the point of origin.

**Fig. 6:**
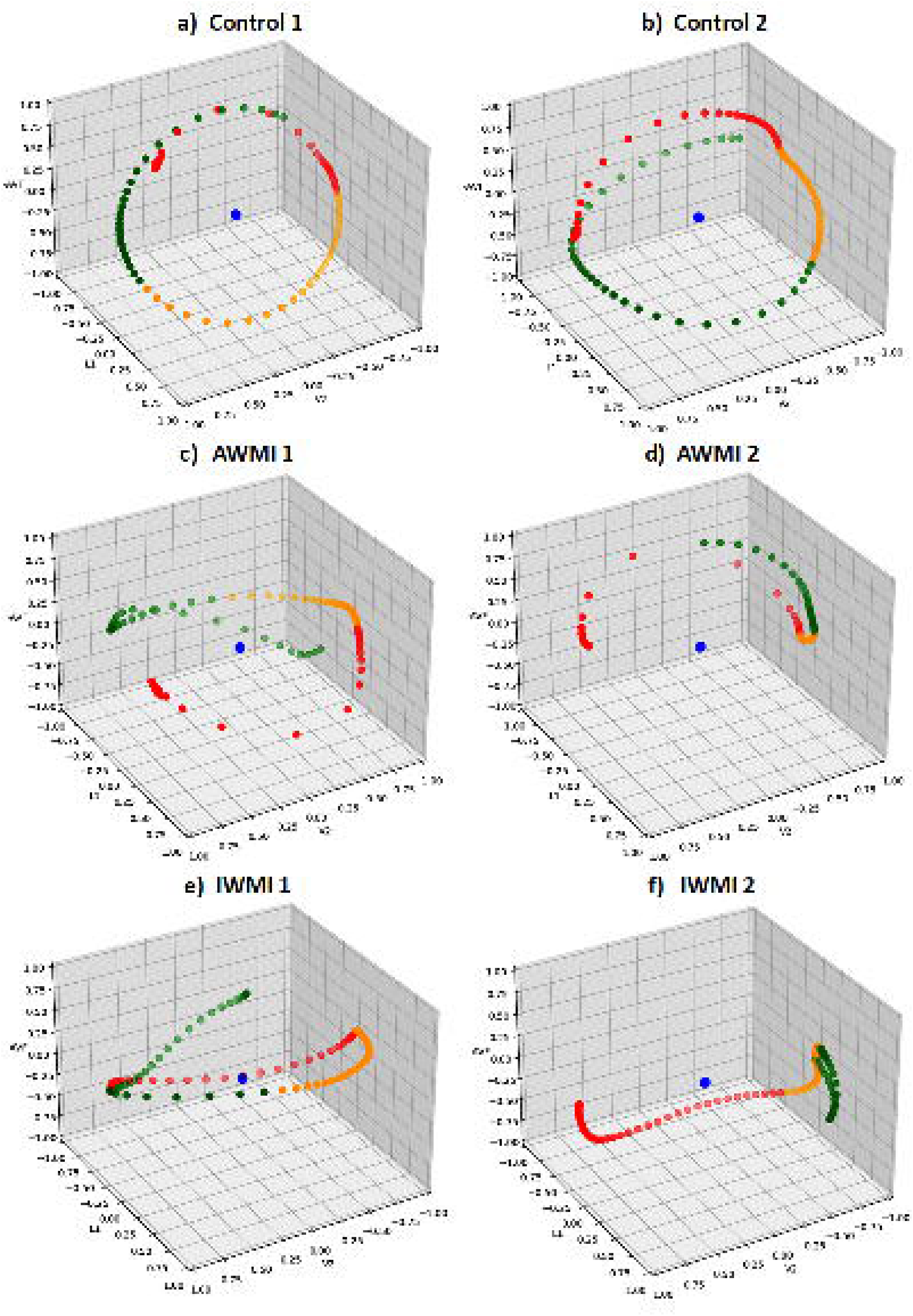
Illustrative representation of Spatial QRS Loop depicted in unit vector system in Control volunteetrs, Patients of AWMI and IWMI with two subjects in each group. The loops are divided into three phases: the early phase is marked in green, the mid-phase in orange, and the final phase in red. The blue dot indicates the point of origin.

As all these figures will be scaled down in the printed version of the article, calibrations and pattens may be challenging to discern. So, their soft copies are also added as supplementary online material which can be viewed on a computer screen and magnified manually to appreciate the details and patterns.

Figures can be freely rotated on the computer screen in order to visualise the VCG loop from different perspectives and spatial positions, including the posterior part of the heart, which cannot be otherwise evaluated directly.

Animated video representations of the QRS-VCG loop of a control and two cases of AWMI and IWMI; which are rotated in the horizontal direction are added as supplementary video materials, for comparative evaluation of visual pattern. In real time experimental evaluation, these loops can be manually rotated in all possible orientations to observe various aspects.

## Validation and discussion

The initial assessment of VCG loop is carried out on the unmodified spatial QRS loops with minimum scale of the reference axes frame in each individual (Fig. 1). This clearly exhibits the distinct spatial dimensions; morphology, contour etc. The illustrated images clearly distinguish between spatial features of VCG loop between control volunteers and AMI, and also within subgroups of AMI. The pathway of the QRS loop varies between control and cases and also between subgroups of cases. The direction of propagation of the spatial QRS loop is also evaluated and it is observed in the illustrated cases and controls that, while the loop is propagating counter clockwise in controls, the directionality shifts to a clockwise pattern in the AWMI case group. In IWMI the morphology of the spatial QRS loop also shows varied patterns in case group; The loop becomes twisted, tortuous, open-ended or other forms in case group instead of a regular counterclockwise closed loop in control volunteers.

Next, we standardized the reference axes to allow us to compare the loop sizes across different individuals. When this was done, the size of the loop became comparable across and within groups. It is clearly visible that the size of the loops is much less in cases of AMI (Fig. 2).

We then analysed the distribution of the VCG loops across the various octants of a cartesian coordinate system with respect to three orthogonal planes. This revealed distinct patterns between healthy individuals and those with AMI, as well as between subtypes of AMI (Fig. 3). For example, in healthy subjects, most of the loop occupied octant 5, while in AWMI, the loops predominantly appeared in octant 6, and in inferior wall myocardial infarction (IWMI), they were mostly in octant 1. This suggests that the electrical current in the heart is altered by myocardial infarction, causing the loops to shift away from the infarcted area.

Evaluation of spatial VCG loop in various cartesian octants of fixed frame of reference representing the distribution of loop morphology, shows the objective dispersion of the loop in various octants with respect to the orthogonal planes (Fig. 4).

Representation of Spatial VCG Loop depicted in unit sphere system shows the exact directional patterns of the vectorcardiography loop in space. By normalising the magnitude component of the vectors, this style of representation provides a method to trace the exact pathway of electrical propagation. When the magnitude component of the vectors is reduced by appropriate transformation the direction component is revealed. This is utilised to map the electrical propagation pathway in each individual subject. This is a novel way of visualization and offers new insights into the electrical activity of the heart.

The unit sphere and unit vector representations vividly depict the point-to-point directional propagation of the vector tips. It is known that with loss of structural and functional properties of the myocardium either locally or in a widespread manner affect the origin, magnitude, direction and propagation of vectors [4]. These unit vector & sphere systems that specifically represent the directionality component of vectors, offer valuable insights regarding the infarct location, altered propagation pattern, extent of the tissue damage, area of myocardial quiescence, stunning, hibernation etc.

These findings support the idea that VCG signals, though complex, offer detailed insights into heart function. However, because the signals are so sensitive to subtle changes, standardizing the analysis remains a challenge. Nevertheless, the visual methods we employed provided useful tools for examining and understanding the QRS loops in both healthy individuals and those with AMI.

## Summary and conclusion

The illustrative methods presented here offer a wide-ranging visual approach to the evaluation and analysis of the 3D VCG loop, each with its unique strengths in envisaging and interpreting the heart’s electrical activity.

The first and second methods (Figs. 1, 2) are highly effective for visualizing and analysing the unmodified electrical activity of the ventricles during the QRS complex of the cardiac cycle. By color-coding the loop into three equal phases—green, orange, and red — the progression of ventricular depolarization can be tracked along with the localization of the early, major and late ventricular vectors. The consistent orientation of the plot ensures that comparisons across different individuals are reliable, making it especially useful for distinguishing between cardiac conditions such as anterior wall myocardial infarction (AWMI) and inferior wall myocardial infarction (IWMI). These conditions often exhibit distinct loop sizes, orientation and morphologies, which are crucial for identifying specific types of myocardial damage and predict outcome.

Building on these methods, the third method (Fig. 3) adds an extra layer of analysis by projecting three planes—Frontal, Sagittal, and Transverse—onto the 3D loop. This allows us to view the loop from multiple angles at once, making it easier to spot subtle differences that may signal various heart problems. This perspective provides a more complete view of the loop’s structure, which is essential for accurate diagnosis and understanding of heart conditions.

The fourth method (Fig. 4) further refines the analysis by color-coding the VCG loop into distinct octants using a variety of colours. This method allows researchers to precisely track the progression of ventricular depolarization within specific spatial segments. It is known that, the propagation of the depolarising current diverts away from the area of infarction, leading to altered pattern of spatial occupation of the loop in various octants. The octant-based approach reveals distinct patterns and variations in vector tip distribution in cases of AWMI and IWMI compared to healthy controls, helping to identify and analyze octant-specific changes within the VCG loop.

Finally, the fifth method (Figs. 5, 6) introduces a unique approach by plotting the QRS loop as a unit vector on a unit sphere to focus solely on the directional movement of the ventricular current. By removing the influence of magnitude data from the unit sphere, it ensures comparability across scales, enabling fair comparisons. The script’s 3D visualization capabilities further facilitate intuitive exploration of data points in relation to the unit sphere, promoting deeper insights and hypothesis generation in scientific investigations.

Unlike the previous methods, which consider both direction and magnitude, this method is particularly useful for identifying infarcts, as it highlights shifts in the current’s original path. Additionally, it provides a means to differentiate AMI subgroups by comparing their QRS vector patterns with those of a control group, offering further insights into specific cardiac conditions.

Together, these four methods provide a robust framework for the comprehensive evaluation of the 3D VCG loop, each contributing to a deeper understanding of cardiac function and the identification of various heart conditions.

## Supporting information

Animated video representations of the QRS-VCG loop of a control volunteer, which are rotated in the horizontal direction

Animated video representations of the QRS-VCG loop of a case of AWMI, which is rotated in the horizontal direction

Animated video representations of the QRS-VCG loop of a case of IWMI, which is rotated in the horizontal direction

## Declaration of competing interest

### Declaration

a. The present manuscript is not under consideration elsewhere.
b. None of the manuscript’s contents have been previously published.
c. All authors have read and approved the manuscript and approved its submission to ***Future Cardiology.*** They also agreed the order of authorship.

### Statements relating to our ethics and integrity policies

We pledge to make wise use of our resources and to be good stewards of financial, capital, and human resources. We operate within the letter and spirit of the law and prescribed policies, and strive to avoid impropriety or conflict of interest.

### Data availability statement

We are ready to make available the relevant research data to improve the transparency of research and accelerate the pace of discovery, so that more research can be independently verified or made reproducible, in future.

### Ethics approval statement

The ethical approval for studies involving human subjects was obtained from the Institutional Ethics Committee of the Institute of Postgraduate Medical Education & Research, Calcutta, India, vide No. IPGME&R/IEC/2021/114 dated 06/02/2021

### Patient consent statement

Written informed consent was obtained from each of the study subjects.

### Clinical trial registration

NA

### Permission to reproduce material from other sources

NA

## Author contributions statement

1. **Ms. Tania Ghosal:**
  a. She made a significant contribution in the conception and study design, including execution & acquisition of data, analysis and interpretation.
  b. She drafted and substantially revised the article.
  c. Have agreed to submit the manuscript in Future Cardiology.
  d. She reviewed and agreed on all versions of the article before submission.
  e. She agreed to take responsibility and be accountable for the contents of the article and to share responsibility to resolve any questions raised about the accuracy or integrity of the published work.
2. **Dr. Subhankar Kumar:**

a. He made a significant contribution in the conception, study design, including acquisition of data, analysis and interpretation.
b. He critically reviewed the article.
c. Have agreed to submit the manuscript in Future Cardiology.
d. He reviewed and agreed on all versions of the article before submission.
e. He agreed to take responsibility and be accountable for the contents of the article and to share responsibility to resolve any questions raised about the accuracy or integrity of the published work
3. **Dr. Imran Ahmed:**

a. He made a significant contribution in the study design, including acquisition of data, analysis and interpretation.
b. He critically reviewed the article.
c. Have agreed to submit the manuscript in Future Cardiology.
d. He reviewed and agreed on all versions of the article before submission.
e. He agreed to take responsibility and be accountable for the contents of the article and to share responsibility to resolve any questions raised about the accuracy or integrity of the published work
4. **Dr. Damodar Prasad Goswami:**

a. He made a significant contribution in the conception, mathematical scheme, study design, analysis and interpretation.
b. He drafted and substantially revised and critically reviewed the article.
c. Have agreed to submit the manuscript in Future Cardiology.
d. He reviewed and agreed on all versions of the article before submission.
e. He agreed to take responsibility and be accountable for the contents of the article and to share responsibility to resolve any questions raised about the accuracy or integrity of the published work
5. **Dr. Anupam Bandyopadhyay:**

a. He made a significant contribution in the study design, analysis and interpretation.
b. He critically reviewed the article.
c. Have agreed to submit the manuscript in Future Cardiology.
d. He reviewed and agreed on all versions of the article before submission.
e. He agreed to take responsibility and be accountable for the contents of the article and to share responsibility to resolve any questions raised about the accuracy or integrity of the published work
6. **Prof. Arnab Sengupta: Corresponding Author**

a. He made a significant contribution in the conception, study design, analysis and interpretation.
b. He drafted, written and substantially revised and critically reviewed the article.
c. Have agreed to submit the manuscript in Future Cardiology.
d. He reviewed and agreed on all versions of the article before submission.
e. He agreed to take responsibility and be accountable for the contents of the article and to share responsibility to resolve any questions raised about the accuracy or integrity of the published work

## Acknowledgment

The Director, Institute of postgraduate medical education & research-SSKM Hospital. 244 AJC Bose Road. Calcutta 700020. India, for permitting us to undertake the present research work and providing all necessary support and logistics.

## Plain Language Summary

The human heart generates electrical signals that control its rhythm and pumping action. Doctors typically record these signals as an electrocardiogram (ECG), but this approach only represents a two-dimensional view. A more detailed view comes from a technique called vectorcardiography (VCG), which traces the heart’s electrical activity in three dimensions, offering a clearer picture of how the heart works.

Our study focuses on visualizing the VCG’s QRS loop, which represents a key phase in the heart’s electrical cycle. We developed five new methods to display the QRS loop:

1. Unmodified VCG Loop: Shows the heart’s natural electrical activity using 3D color-coded loops.
2. Fixed-Scale Loop: Standardizes the size of the loop, making comparisons between individuals easier.
3. Octant Analysis: Divides the 3D space into eight sections, helping to spot unusual patterns linked to heart damage.
4. Unit Sphere Mapping: Highlights the direction of electrical signals by projecting the loop onto a standard unit sphere, removing the effect of signal strength.
5. Unit vector mapping: Represents the direction of electrical signal of the loop keeping aside the magnitude of the vector.

We applied these techniques to study people with normal heart function and those with heart attacks affecting the front or bottom walls of the heart. The visual representations revealed clear differences in loop size, direction, and shape, helping us understand how heart damage alters electrical signals.

Our work demonstrates how advanced VCG visualization can enhance heart condition diagnosis and treatment planning. By making heart signal analysis more precise, we hope to improve how doctors detect and monitor heart problems.

## Article Highlights

▪ Advanced Visualization of Heart Signals: Introduced four innovative methods for visualizing the QRS loop in 3D-vectorcardiograms (VCGs).
▪ Enhanced Diagnostic Insights: Demonstrated how these methods provide deeper insights into heart function, especially in conditions like anterior and inferior wall myocardial infarctions.
▪ Standardized Comparisons: Developed a fixed-scale VCG representation enabling direct comparisons across individuals and heart conditions.
▪ Spatial Distribution Analysis: Proposed octant-based VCG analysis to detect spatial abnormalities caused by heart damage.
▪ Directional Focus Using Unit Sphere: Separated magnitude and direction of heart signals using a unit sphere model for clearer signal path analysis.
▪ Unit vector mapping: Indicates the direction of the electrical signal in the loop keeping aside the magnitude of the vector.
▪ Clinical and Research Applications: Showed the potential of these visualization methods for advancing both clinical diagnostics and cardiac research

